# Assessment of accuracy of detection dog signaling behavior for the diagnosis of SARS-CoV-2 infection: A Canadian study

**DOI:** 10.64898/2026.03.04.26347154

**Authors:** Fiston Ikwa Ndol Mbutiwi, Colombe Otis, Ian Schiller, Mathieu LaChance, Laurie Martin, Ali Jammal, Amaka Odita, Nancy Agbaje, Aisha Khatib, Nandini Dendukuri, Hala Tamim, Eric Troncy, Hélène Carabin

**Affiliations:** Faculté de Médecine Vétérinaire, Université de Montréal, Saint-Hyacinthe, Québec, Canada; Groupe de Recherche en Épidémiologie des Zoonoses et Santé Publique (GREZOSP), Saint-Hyacinthe, Québec, Canada; Centre de Recherche en Santé Publique (CReSP), Montréal, Québec, Canada; Faculty of Medicine, University of Kikwit, Kikwit, Kwilu, Democratic Republic of the Congo; Groupe de Recherche en Pharmacologie Animale du Québec (GREPAQ), Université de Montréal, Saint-Hyacinthe, Québec, Canada; Research Institute of the McGill University Health Centre (RI-MUHC), Montreal, Quebec, Canada; University College Cork, Cork, Ireland; Boston University, Boston, Massachusetts, United States of America; Clinical program, Health Ontario, Toronto, Ontario, Canada; Department of Family & Community Medicine, University of Toronto, Toronto, Ontario, Canada; Unity Health Toronto (UHT), Toronto, Ontario, Canada; York University, Toronto, Ontario, Canada; École de Santé Publique, Université de Montréal, Montréal, Québec, Canada

**Keywords:** detection dogs, sniffer dogs, COVID-19, volatile organic compounds, diagnostic test, Bayesian latent class models

## Abstract

**Background:** Dogs trained to metabolomics detection can identify pathological changes through their refined smelling sense. During the COVID-19 pandemic, studies worldwide evaluated Detection Dog Signaling Behavior (DDSB) for SARS-CoV-2. However, most statistical approaches failed to account for key sources of bias, potentially distorting performance estimates. This study aimed to estimate DDSB accuracy for SARS-CoV-2 infection in a Canadian population while assessing the impact of selected sources of bias on performance estimates.

**Methods:** Participants attending the COVID-19 assessment clinic at St. Joseph’s Health Centre, Toronto, were recruited between October and December 2021. Each provided a nasopharyngeal swab for reverse transcription-polymerase chain reaction (RT-PCR) testing and three sweat samples for canine detection. Three dogs were trained to detect SARS-CoV-2 in sweat samples. Validation sessions were video recorded and independently reviewed by two blinded observers. DDSB diagnostic accuracy was estimated against RT-PCR, evaluating the impact of ignoring its imperfect accuracy and repeated sniffing of the same samples during validation.

**Results:** Among 2,358 participants (mean age: 34.7 years; 55.7% female), 437 contributed to training. Validation tests included 146 unique participants (25 RT-PCR positive, 121 negative). Assuming RT-PCR was imperfect, DDSB posterior median sensitivity ranged from 67% (95% credible interval [CrI]: 29%–97%) to 78% (95% CrI: 41%–99%), and specificity from 67% (95% CrI: 53%–79%) to 77% (95% CrI: 65%–87%) across the three dogs. Assuming RT-PCR was perfect, sensitivity decreased by 7.9% to 9%, while specificity remained unchanged. Including repeated positive samples without adjustment did not affect specificity estimates but overestimated sensitivity by 7.9% to 13.4% (imperfect RT-PCR) and 11.4% to 18.3% (perfect RT-PCR).

**Conclusion:** DDSB shows potential as a non-invasive screening tool for SARS-CoV-2 infection. Our results highlight the challenges of designing such studies and the need for standardized training and validation procedures to ensure the reliability and validity of DDSB.

## 1. Introduction

The fast emergence and global spread of Severe Acute Respiratory Syndrome Coronavirus 2 (SARS-CoV-2) prompted the unprecedented rapid development and deployment of new diagnostic and screening tests to support clinicians and public health authorities in managing the pandemic response [1,2].

While reverse transcription-polymerase chain reaction (RT-PCR) tests are considered highly accurate, with pooled sensitivity estimated at 96% (95% Confidence Interval [CI]: 93%−98%) and pooled specificity at 100% (95% CI: 98%–100%)) [3], their use in screening settings raised practical challenges. These included invasive testing, high costs, delays in obtaining results, the need for qualified personnel, and limited availability of testing reagents, all contributing to significant bottlenecks as the pandemic spread [4–6]. Rapid antigen tests, which detect specific SARS-CoV-2 antigens in host tissues or body fluids, have shown excellent specificity, with pooled estimates ≥ 95%. However, their sensitivity was lower, with pooled estimates ranging from 62% to 70% overall, and between 55% and 71% in analyses excluding outliers [7].

In this context, several researchers [8,9] have investigated the use of detection dog signaling behavior (DDSB) as a screening tool for identifying SARS-CoV-2 infection. The human body produces a diverse array of volatile (and semi-volatile) compounds, varying with individual factors such as age, gender, diet, genetics, and pathophysiological status. These volatile organic compounds (VOCs) constitute a unique volatilome for each individual [10]. Therefore, pathological processes can alter this biochemical profile, influencing body odor by either the production of specific compounds, or more surely modifying the overall profile [11] to create a distinctive “olfactory fingerprint”. Recently, the VOCs profile of patients diagnosed with SARS-CoV-2 was identified by breath [12], blood [13,14], and sweat [15] analyses. Indeed, the SARS-CoV-2 specific volatilome has been recognized by dogs in samples from breath, saliva, sweat, urine, feces, skin, and blood [10,16–19]. In all cases, the dog’s behavior must be interpreted by a human, *i.e.* the testing modality is human observation of the DDSB. A systematic review reported very good diagnostic performance of DDSB for SARS-CoV-2, with sensitivity estimates ranging from 82% and 97% and specificity from 83% and 100% in six high-quality studies out of 27 published studies [9]. Such promising results suggest that DDSB offers an alternative to RT-PCR and rapid antigen tests for early, rapid, inexpensive, reliable, ecological, and non-invasive screening of large populations in settings such as airports, schools, universities, workplaces, and events.

In Canada, few studies have evaluated the validity of using DDSB for COVID-19. Two studies conducted in Vancouver, both employing the same two dogs, reported sensitivity estimates of 100%, with specificity ranging from 83% to 100% among long-term care residents [20], and from 93% to 94% in the general population [21]. However, these studies [20,21], along with many others worldwide [22–39], evaluated DDSB performance using RT-PCR as the reference standard, without accounting for its potential imperfection in classifying SARS-CoV-2 infection cases [40–44]. Moreover, substantial heterogeneity, and even complete absence of reporting, persisted in key methodological areas, including participant selection, detection dog selection and training, observer selection, specimen type, sample collection, storage and handling, experimental settings and study design, training and validation protocols, criteria for transitioning from training to validation, and results reporting [45]. This undermines the reproducibility and complicates assessment of bias in DDSB accuracy estimates. Consequently, interpreting results of individual studies, and synthesizing results across studies remain challenging.

This study primarily aimed to estimate the performance of DDSB in screening for SARS-CoV-2 infection within a Canadian population, while accounting for the imperfect accuracy of RT-PCR through Bayesian latent class models (BLCM). A secondary objective was to assess the impact of repeated sniffing of the same samples by the dogs during the validation phase on DDSB performance estimates.

## 2. Materials and methods

The reporting of this diagnostic study was conducted in accordance with the Standards for the Reporting of Diagnostic accuracy studies that use BLCM (STARD-BLCM) checklist (S1 Appendix) [46]. All procedures were carried out in compliance with the principles of the Declaration of Helsinki, the requirements of ICH GCP Harmonised Tripartite Guideline Topic E6 and Directives 2001 / 20 / EC and 2005 / 28 / EC of the European Parliament, the ethical practices established by Unity Health Toronto (UHT), and followed Good Clinical Practice (GCP) guidelines. Written ethical approval for the collection of samples and the gathering of information from recruited participants was obtained from the Research Ethics Board of UHT (University of Toronto; REB #21-236C), and the Research Ethics Committee in Science and Health (Université de Montréal; #2021-1206). Each participant provided verbal informed consent, which was documented by signing a consent form confirming their verbal agreement. As for the use of animals, ethical approval was granted by the Comité d’Éthique de l’Utilisation des Animaux at the Faculty of Veterinary Medicine of the Université de Montréal (#21-Rech-2165).

### 2.1. Participants

Eligible participants were English-speaking women and men aged 18 or older, or minors with the consent of a legal guardian, visiting the COVID-19 Assessment Clinic at St. Joseph’s Health Centre in Toronto, who agreed to provide sweat samples on the same day they gave nasopharyngeal swab samples for a RT-PCR test to detect SARS-CoV-2. Individuals with clinical contraindications, as determined by the attending physician, were not enrolled in the study. Participation was voluntary and anonymous, with recruitment conducted consecutively from October 12 to December 20, 2021, a period coinciding with the emergence of the Omicron variant. After providing informed consent, participants were administered a short verbal questionnaire to collect clinical information, including age, sex, ethnicity, smoking status, presence of COVID-19-related symptoms, COVID-19 vaccination status and brand received, when applicable.

### 2.2. Sweat samples collection and preparation

Sweat samples were collected with the help of a standardized operating procedure (SOP), from a designated site of Unity Health Toronto (St. Joseph’s Health Centre) by a clinical staff consisting of doctors, nurses, research assistants, and clinical research agents, all of whom had undergone specific training in sample collection and handling. Each participant was provided with a sweat sample collection kit that included two pairs of nitrile gloves, two sterile gauzes for sampling sweat from the face (forehead, cheeks), throat, and neck, four dental cotton gauzes for both axillary sweat samples, and three anti-UV sterile pill bottles (one for the gauzes soaked with sweat from the face, throat, and neck, one for the gauzes from the left armpit, and one for the gauzes from the right armpit). The participant was asked to place two dental gauzes underneath each armpit for 30 seconds, then use one sterile gauze to rub the forehead, both cheeks, and the throat and neck area. These gauzes were then placed in the corresponding coded pill bottles. After sample collection, participants were asked to remove their nitrile gloves, dispose of them in a red biological waste bag, and disinfect their hands with alcohol gel.

The sweat samples were labeled with an anonymized identification code linked to the corresponding patient’s RT-PCR result. The coded pill bottles were stored together in a bag, which was also labeled with a code. The sealed bags were then stored in a cardboard box and once full, sent to the study site, where they were kept at a temperature between 0°C and 10°C until use. All surfaces were cleaned with 96% isopropyl alcohol before and after processing the samples. Preventive measures against COVID-19 were strictly followed for clinical staff, including the use of gloves and N95 masks during sample collection and handling, frequent hand washing and facility disinfection, as well as weekly COVID-19 testing.

### 2.3. Implementation of RT-PCR test

RT-PCR test was the reference test for SARS-CoV-2 infection detection. It was processed at the St. Michael’s Hospital Microbiology department using participants’ nasopharyngeal swab samples and results reported to patient’s clinical charts. A cycle threshold (Ct) value < 40 was used to define RT-PCR positivity [47].

### 2.4. Implementation of DDSB test

The DDSB was the index test and consisted of the observer’s judgment of whether a sample was positive or negative for SARS-CoV-2, based on the behavior of a dog exposed to that sample, as observed through a muted video recording of a validation session.

#### 2.4.1. Dogs and handler

Three detection dogs were included in the study: two Labrador Retrievers, one male (Dog 1) and one female (Dog 2), and one female German Shepherd (Dog 3). Dog 1 had no prior experience in scent detection (“green” dog), whereas Dogs 2 and 3 had previous experience detecting bed bugs. The dogs lived at the study site throughout the study period and performed only tasks related to the study. A single dog handler, the owner of all three dogs and experienced in canine bed bug detection, led the dogs through all phases of the study.

#### 2.4.2. Training phase

The dogs were trained to detect specific VOCs associated with SARS-CoV-2 infection from participants’ sweat samples using a medical odor detection training program elaborated by researchers from the Faculty of Veterinary Medicine of the Université de Montréal (E.T. and C.O.), in collaboration with several experts in canine training and odorology, in particular of the Nosaïs Institute (Paris, France) [48], formalized as a SOP (available upon request from the corresponding author). The six-steps training phase lasted approximately 35 days, between April and June 2022, and therefore used sweat samples collected 4 to 6 months earlier, a delay due to administrative and operational constraints. Training took place in a decommissioned school building, which was temperature-controlled with air conditioning and converted into a dog training center. Sessions were supervised by a research assistant (A.J. and/or C.O.) and overseen by a Professor (H.T. and/or E.T.) to ensure compliance with training protocols. No video recordings were made of the training sessions. Briefly, the six steps consisted in 1) Exposure to sniffing and marking (the dog is used to the system of sniffing and marking, with the assistance of a familiar toy, rewarding with clicker use); 2) Impregnation to and marking of positive samples (memorizing the COVID-19 sample odor with rewarding of the dog when it marks the container which includes the positive sample, and all the remaining containers in the line-up are empty); 3) Learning the line-up work (all containers, but one occasional with a positive sample, in the line-up are empty and the dog is rewarded with its toy when it sniffs each of the olfaction containers); 4) Introduction of mock neutral samples (empty containers are replaced by mocks, *i.e.*, containers containing the whole material with sterile gazes; the dog is rewarded when it marks the positive sample); 5) Introduction of negative samples and of the negative mark mat (COVID-19 negative samples are introduced into the line-up which contains one occasional positive sample, one or two mock(s), and negative sample(s)); 6) Introduction of several positive samples and of the all negative mat. Only when a step had been successfully completed was the dog allowed to move to the next step of training. The dogs were trained to sit or look at their handler in front of a hole containing a positive sample, and not to exhibit these behaviors in front of a hole containing a negative sample. If all samples were negative, the dogs were trained to sit on a mat placed away from the sniffing station.

During steps 1 to 4, the samples were presented to the dogs using containers held with microphone holders. From the Step 5 of the training, and during the validation phase, the samples were presented to the dogs using an aluminum-mage sniffing station, which allowed them to potentially sniff up to six (successive) samples in a short period of time (Fig 1). Neither the dog nor the handler had access to the sample placement area located at the back of the station. The samples were contained in pill bottles, which were first inserted into a rack before their lids were opened. The rack was then inserted horizontally behind the main frame, with each bottle fitting tightly into its attributed hole without any gaps. The three dogs took turns sniffing the same samples multiple times, and the sniffing station was cleaned with 96% isopropyl alcohol before and after each dog’s attempt.

**Fig 1.**
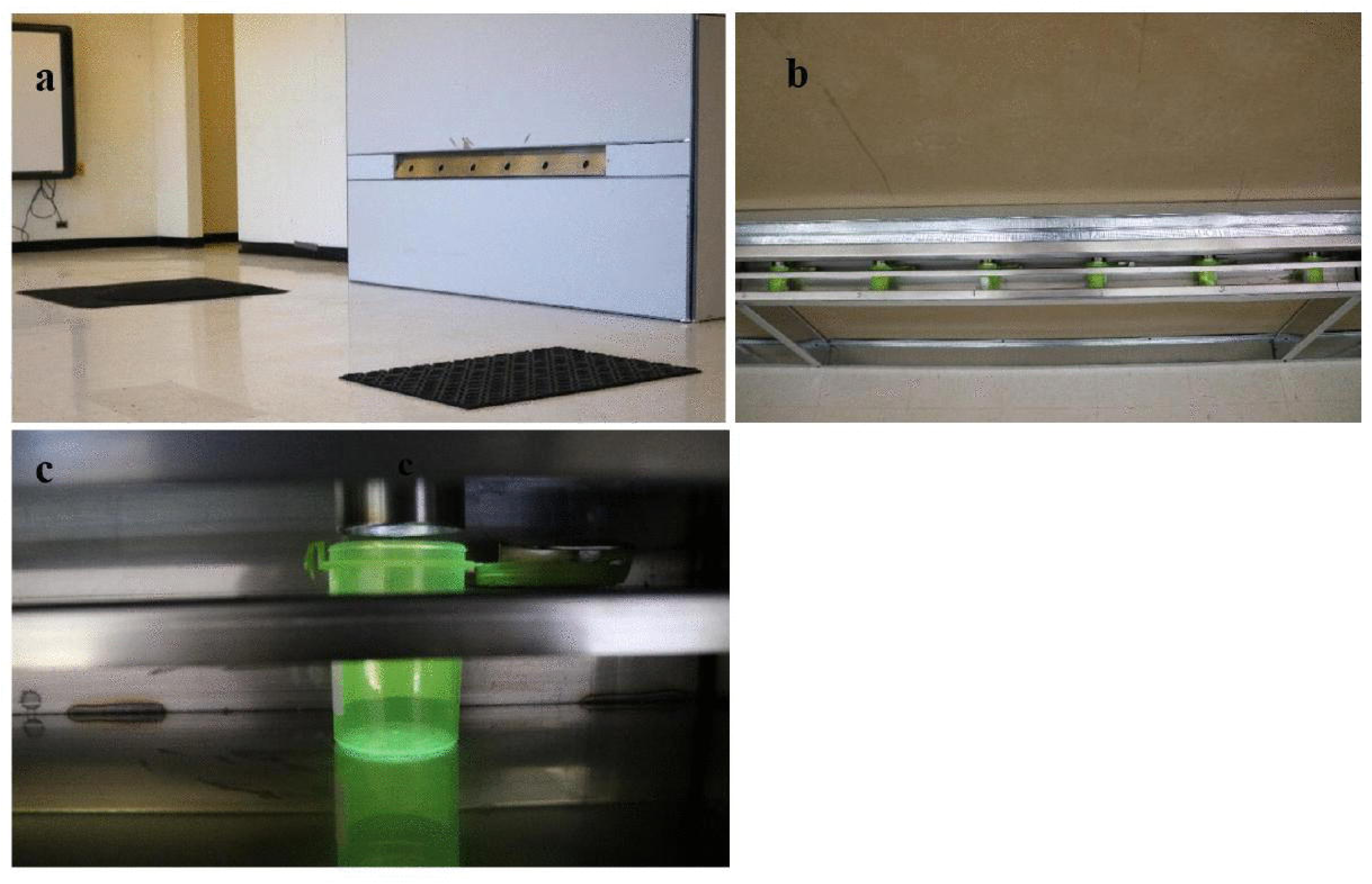
Sniffing station used from step 5 of the training to the validation phase. Front (**a**) and rear (**b**) views of the sniffing station, with a close-up of an open pill bottle next to a hole in the back of the rack (**c**).

Retained positive samples for Steps 2 to 5 (Ct values < 34, n = 60 individuals, 180 samples) were largely from symptomatic patients (78%), non-smokers (92%), without respiratory comorbidities (100%). The negative samples for Step 5 were those from individuals that were non-smokers (except for one vaccinated smoking marijuana), asymptomatic and without comorbidities either unvaccinated (n= 52) or vaccinated (n=213). In step 6, the dog was trained on combinations of positives and negatives and by using samples from individuals with a combination of characteristics regarding their comorbidities, age, smoking and vaccination status as well as including samples taken from the forehead and neck. Step 6 was done using samples from 12 positive and 100 negative individuals. The placement of positive and negative samples on the rack was randomized; however, both the handler and the research assistant were aware of the RT-PCR result associated with each sample, during the training.

Training proceeded until the handler was confident in the dogs’ ability to identify neutral, negative and positive samples. Subsequently in step 6, the dogs were exposed to 20 racks containing 3 samples each (*i.e.*, 60 samples), at a rhythm of 5 racks over a 45-minute period, with each rack containing 0, 1, or 2 positive samples. Each of the three dogs surpassed the 90% correct detection threshold (positive and negative samples), marking the completion of the training phase.

#### 2.4.3. Validation setup

The validation phase was planned to be conducted in two successive rounds in the same setting as the training phase. The first validation round was intended to evaluate the performance of the dogs with samples from individuals they had never sniffed before, as recommended [49]. In this round, each sample was meant to be sniffed by one dog only, since the concentration of the VOC is expected to decrease each time the container with the sample is opened, giving the dog that sniffed the sample first an advantage. The second validation round was planned to include repeated sniffing of the same samples, such that a single sample could be sniffed by multiple dogs and the same sample could be sniffed more than once by the same dog, allowing the assessment of both between-dog and within-dog repeatability. To familiarize the handler with blinded conditions whereas s/he was unaware of both the number and location of positive and negative samples, it was decided to add preliminary sessions to the validation phase. The preliminary sessions included some sample duplications, as we sought to preserve as many un-sniffed individuals as possible for the subsequent validation round. Specifically, dogs were exposed to new samples from certain individuals they had previously sniffed during training, and some samples were presented to multiple dogs or sniffed more than once by the same dog. Due to unforeseen circumstances, the validation phase was prematurely halted after five days. Hence, the data available for analyses included those from the preliminary sessions and part of the first validation round.

All tests were conducted under blinded, randomized, and controlled conditions, supervised by a research assistant. Only the research assistant, who was responsible for placing the sweat samples on the rack, knew the distribution of positive and negative samples; however, he had no contact or interaction with the handler during the testing sessions. The test sessions were recorded using a web platform specifically developed for olfactory detection with dogs. Two independent observers (M.L and L.M) blindly reviewed each video recording outside of the testing sessions and classified each sample as either positive or negative based on the dog’s behavior, as mentioned above. Videos were reviewed without sound to remove any potential impact of the trainer’s verbal responses to the dog’s reactions.

### 2.5. Statistical Analysis

A flow chart described how the study design and sample size for the statistical models was determined (Fig 2). The agreement between both observers’ recordings of each dog’s response was examined using a two-by-two table. Since there was negligible discrepancy between them, only the data from one of the observers (Observer 1) were used in the statistical models to avoid problems due to multicollinearity.

**Fig 2.**
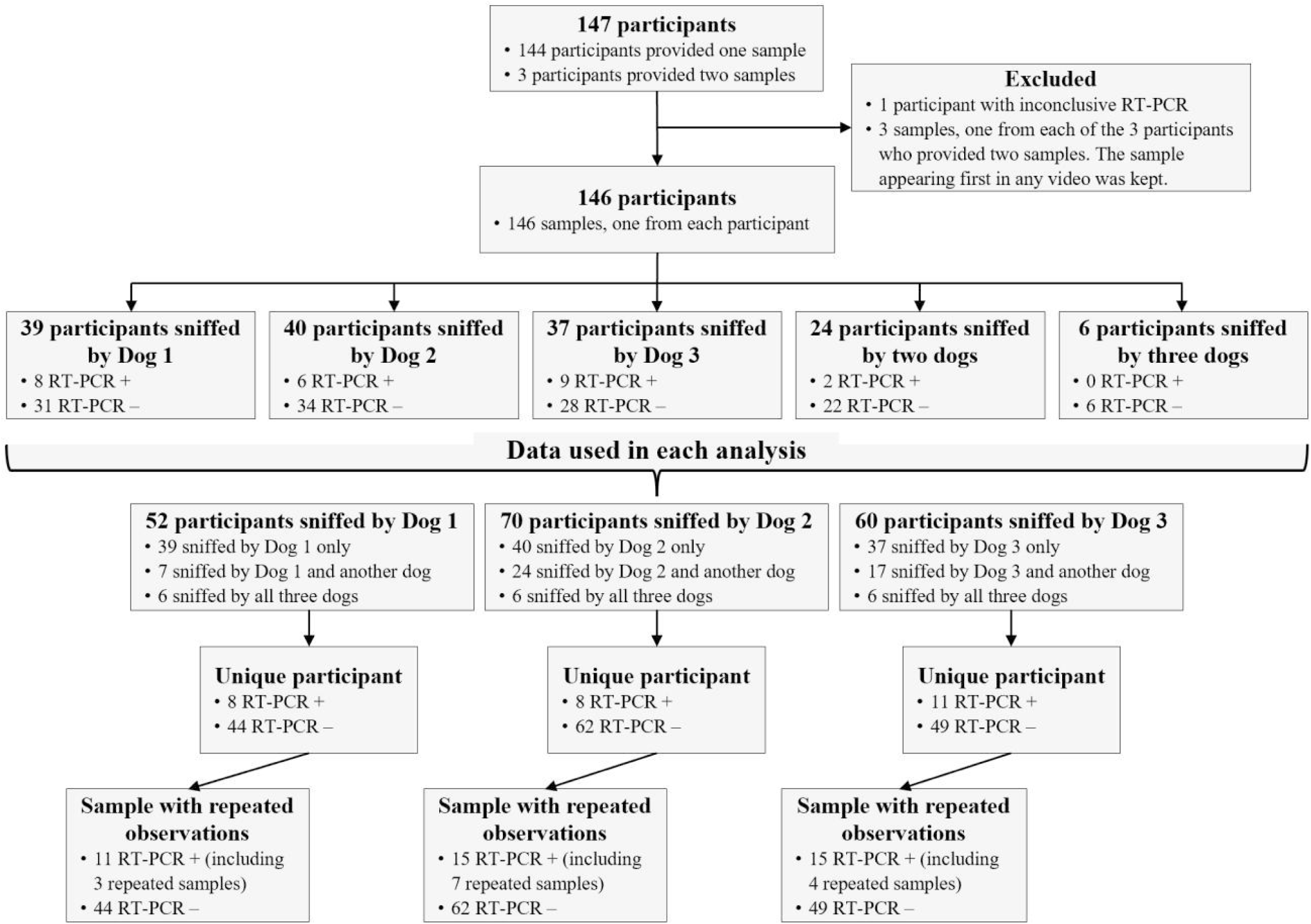
Participants and samples sniffed by each dog during the validation phase, with SARS-CoV-2 RT-PCR results. RT-PCR, reverse transcription-polymerase chain reaction.

In the primary analysis, the diagnostic accuracy of DDSB in identifying COVID-19 was evaluated separately for each dog by comparison to the RT-PCR as the perfect reference test. For each subject, we used only the first sample sniffed by a given dog during the validation. The sensitivity and specificity were calculated based on comparison to the RT-PCR results. A Bayesian approach was used for inference, with non-informative Beta(1,1) prior distributions.

To estimate the bias resulting from treating RT-PCR as perfect, we allowed for informative prior distributions over RT-PCR sensitivity (median: 94.5% (95% confidence interval (CI): 85.1% to 98.0%) represented as Beta(53.1, 4.0)) and RT-PCR specificity (median: 98.4% (95% CI: 86.7% to 99.8%) represented as Beta(29.2, 1.2)) [50,51]. We assumed that detection dogs were independent of RT-PCR conditional on the COVID-19 status. To estimate the bias that would result when the same sample was presented to the same dog more than once during testing, we re-ran the analysis above including the repeated samples (Fig 2).

Analyses were performed using the R statistical software version 4.2.3 [52]. All models were implemented with the rjags package [53]. The model is provided in the supporting information (S2 Appendix). Posterior summaries for sensitivity, specificity, and prevalence of COVID-19 were reported as median estimates with 95% CrIs.

## 3. Results

A total of 2,358 individuals aged 2 to 84 years (mean age: 34.7 years; standard deviation [SD]: 16.6 years) were recruited, of whom 55.7% were female, 71.9% identified as Caucasian, and 25% were smokers. Three-quarters (76.7%) of the recruited subjects reported COVID-19-related symptoms, predominantly respiratory symptoms (90.4%) and fatigue (58.1%). Additionally, 85.2% of the participants had received at least two doses of a COVID-19 vaccine, mostly of the Pfizer brand, and 5.1% had a positive RT-PCR result for COVID-19 (Table 1). The number of subjects whose samples were used in the training phase was 437 (72 RT-PCR positive and 365 RT-PCR negative). Their characteristics were generally consistent with those of the whole sample, except for a lower proportion of smokers (9.4%) and a lower proportion of symptomatic participants (33.4%) (Table 1).

**Table 1.** Participant characteristics in the enrolled, training, and validation samples.

|  | Enrolled sample<br>(n = 2,358) | Training sample<br>(n = 437) | Validation sample<br>(n = 146) |
| --- | --- | --- | --- |
| Age, mean (SD), y | 34.7 (16.6) | 32.3 (15.7) | 35.8 (15.5) |
| Median (IQR), y | 34 (24 – 45) | 32 (20 – 44) | 34 (25 – 45) |
| Gender, n (%) |  |  |  |
| Female | 1,314 (55.7) | 238 (54.5) | 79 (54.1) |
| Male | 1,043 (44.2) | 199 (45.5) | 67 (45.9) |
| Other | 1 (0.0) | 0 (0.0) | 0 (0.0) |
| Ethnicity, n (%) |  |  |  |
| Caucasian, | 1,693 (71.9) | 311 (71.2) | 103 (70.6) |
| Southeast Asian | 137 (5.8) | 25 (5.7) | 4 (2.7) |
| Hispanic | 72 (3.1) | 13 (3.0) | 4 (2.7) |
| Black | 68 (2.9) | 13 (3.0) | 9 (6.2) |
| Indigenous/First Nations | 12 (0.5) | 2 (0.5) | 1 (0.7) |
| Other | 373 (15.8) | 73 (16.7) | 25 (17.1) |
| Smoking status, n (%) |  |  |  |
| Non-smokers | 1,769 (75.0) | 396 (90.6) | 106 (72.6) |
| Smokers | 589 (25.0) | 41 (9.4) | 40 (27.4) |
| COVID-19 symptom status, n (%) |  |  |  |
| Asymptomatic | 550 (23.3) | 291 (66.6) | 19 (13.0) |
| Symptomatic | 1,808 (76.7) | 146 (33.4) | 127 (87.0) |
| Types of symptoms, n (%) | (n = 1,808) | (n = 146) | (n = 127) |
| Respiratory symptoms | 1,635 (90.4) | 136 (93.2) | 121 (95.3) |
| Fatigue | 1,051 (58.1) | 89 (61.0) | 70 (55.1) |
| Pain signs | 405 (22.4) | 46 (31.5) | 32 (25.2) |
| Digestive symptoms | 302 (16.7) | 18 (12.3) | 20 (15.8) |
| Fever | 281 (15.5) | 23 (15.8) | 18 (14.2) |
| Loss smell | 47 (2.6) | 10 (6.9) | 8 (6.3) |
| COVID-19 vaccination status, n (%) |  |  |  |
| No COVID-19 vaccination | 299 (12.7) | 78 (17.9) | 16 (11.0) |
| Received 1st dose | 45 (1.9) | 5 (1.1) | 0 (0.0) |
| Received both doses | 1,895 (80.4) | 333 (76.2) | 119 (81.5) |
| Received three doses | 114 (4.8) | 20 (4.6) | 11 (7.5) |
| Missing | 5 (0.2) | 1 (0.2) | 0 (0.0) |
| Vaccine brand for 1st and/or 2nd doses, n (%) | (n = 2,055) | (n = 358) | (n = 130) |
| Pfizer | 1,168 (56.8) | 209 (58.4) | 68 (52.3) |
| Moderna | 247 (12.0) | 38 (10.6) | 22 (16.9) |
| AZ | 80 (3.9) | 13 (3.6) | 7 (5.4) |
| Mix-Pfizer/Moderna | 351 (17.1) | 68 (19.0) | 23 (17.7) |
| Mix-AZ/Moderna | 121 (5.9) | 19 (5.3) | 5 (3.9) |
| Mix-AZ/Pfizer | 80 (3.9) | 11 (3.1) | 5 (3.9) |
| Other | 8 (0.4) | 0 (0.0) | 0 (0.0) |
| COVID-19 RT-PCR result, n (%) |  |  |  |
| Negative | 2,220 (94.2) | 365 (83.5) | 121 (82.9) |
| Positive | 121 (5.1) | 72 (16.5) | 25 (17.1) |
| Inconclusive | 17 (0.7) | 0 (0.0) | 0 (0.0) |

SD, standard deviation; IQR, interquartile range; COVID-19, coronavirus disease 2019; AZ,
Astra Zeneca; RT-PCR, reverse transcription-polymerase chain reaction.

Validation tests were performed on 146 samples from 146 unique participants (25 RT-PCR positive not yet sniffed by the dogs and 121 randomly selected RT-PCR negative) (Table 1). Each dog evaluated a subset of subject samples, with only 6 out of 146 samples being evaluated by all three dogs. A total of 12 unused positive samples and 45 unused negative samples from participants previously sniffed during training were used during the validation phase. Additionally, 14 RT-PCR positive samples (3 for Dog 1, 7 for Dog 2, and 4 for Dog 3) were presented more than once to the dogs across the validation sessions (Fig 2).

Table 2 presents the diagnostic performance of DDSB based on the primary analysis with unique participation, as well as the sensitivity analysis adjusting for the imperfect accuracy of RT-PCR. When RT-PCR was assumed to be an imperfect reference test, the estimated posterior median sensitivity ranged from 67% (95% CrI: 29%–97%) to 78% (95% CrI: 41%–99%) across the three dogs, and specificity ranged from 67% (95% CrI: 53%–79%) to 77% (95% CrI: 65%–87%). When RT-PCR was assumed to be a perfect reference test, sensitivity estimates decreased by 7.9% to 9.0%, while specificity remained unchanged.

**Table 2.**
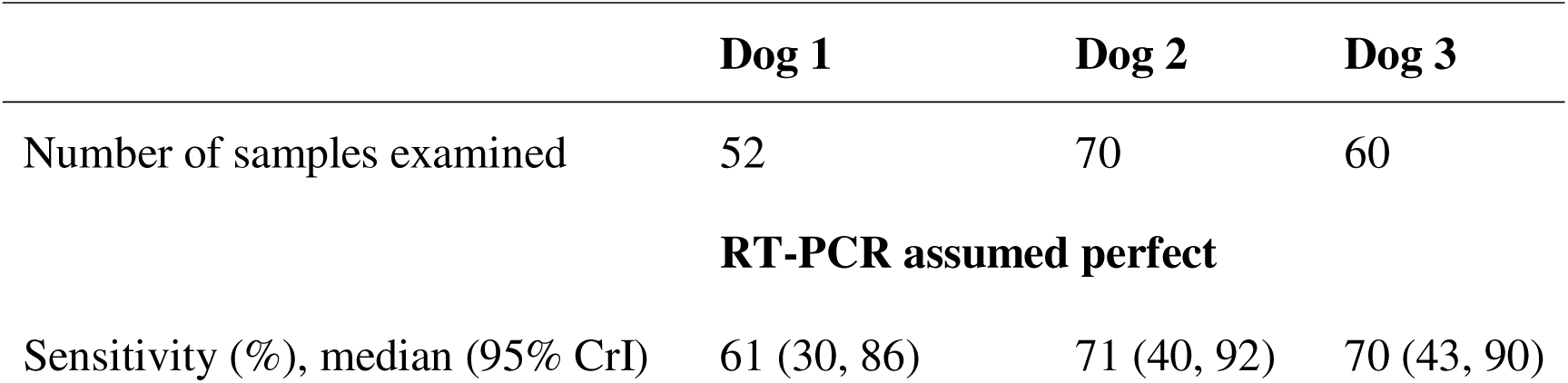
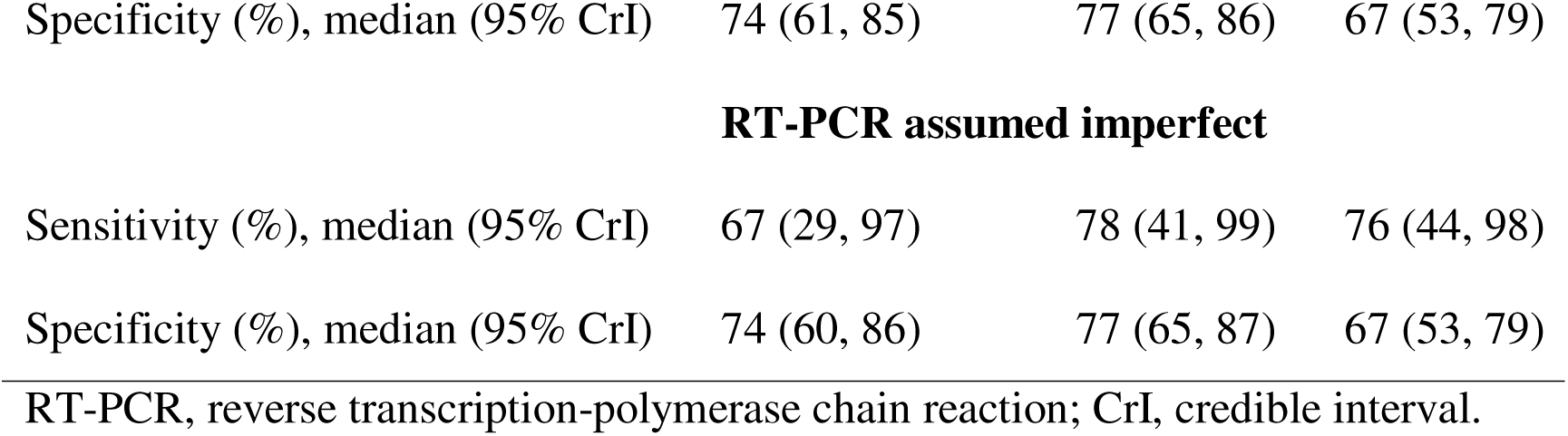
Diagnostic accuracy of human observation of three trained detection dogs in identifying COVID-19 compared to RT-PCR as the reference test. Unique participation of sample per dog.

|  | Dog 1 | Dog 2 | Dog 3 |
| --- | --- | --- | --- |
| Number of samples examined | 52 | 70 | 60 |
| **RT-PCR assumed perfect** |  |  |  |
| Sensitivity (%), median (95% CrI) | 61 (30, 86) | 71 (40, 92) | 70 (43, 90) |
| Specificity (%), median (95% CrI) | 74 (61, 85) | 77 (65, 86) | 67 (53, 79) |
| **RT-PCR assumed imperfect** |  |  |  |
| Sensitivity (%), median (95% CrI) | 67 (29, 97) | 78 (41, 99) | 76 (44, 98) |
| Specificity (%), median (95% CrI) | 74 (60, 86) | 77 (65, 87) | 67 (53, 79) |

RT-PCR, reverse transcription-polymerase chain reaction; CrI, credible interval.

When repeated samples were retained in the analysis (Table 3), sensitivity estimates ranged from 76% (95% CrI: 45%–98%) to 88% (95% CrI: 64%–99%), and specificity estimates ranged from 68% (95% CrI: 54%–80%) to 78% (95% CrI: 66%–88%), under the assumption that RT-PCR was an imperfect reference test. When RT-PCR was assumed to be a perfect reference test, sensitivity estimates decreased by 4.5% to 7.9%. As in the analysis with unique participants, specificity estimates remained stable.

**Table 3.**
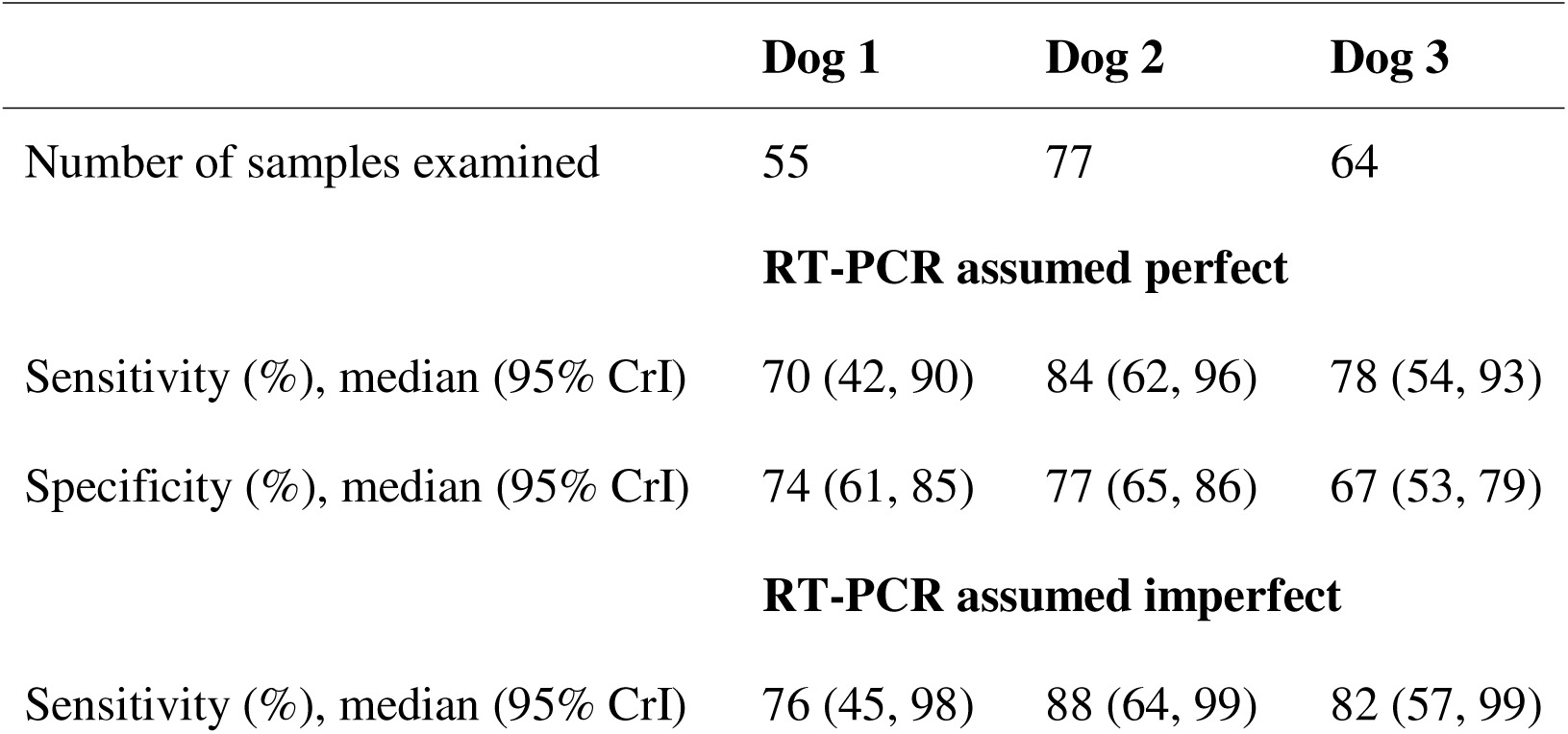
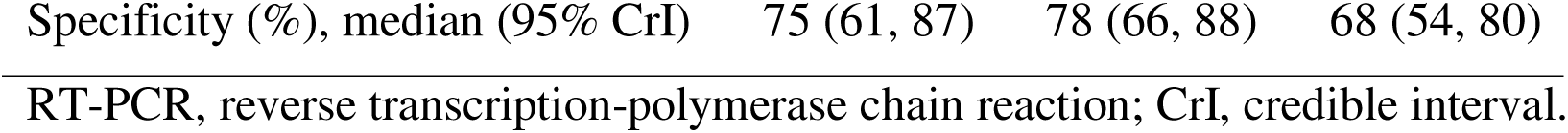
Diagnostic accuracy of human observation of three trained detection dogs in identifying COVID-19 compared to RT-PCR as the reference test. Including same samples repeated in testing per dog.

The comparison of analyses that included and excluded repeated observations shows that ignoring repeated samples did not affect the estimated specificity of DDSB as only positive PCR samples were repeated. Including these samples resulted in an increase in sensitivity estimates by 7.9% to 13.4% when RT-PCR was assumed to be an imperfect reference test, and by 11.4% to 18.3% when RT-PCR was considered a perfect reference test.

We present in the supporting information the diagnostic performance of DDSB as assessed by the two observers (S1 Table), as well as the diagnostic accuracy of RT-PCR when assumed to be imperfect (S2 Table).

## 4. Discussion

The present study focused on estimating the accuracy of observing the response of detection dogs in identifying SARS-CoV-2 infection and explored the impact of selected sources of bias on performance estimates. Previous published studies have evaluated the use of DDSB for diagnosing SARS-CoV-2, either through direct sniffing of individuals [54,55] or through various biological specimens such as sweat, breath, gargle, saliva or urine [8,21]. However, substantial methodological heterogeneity across these studies may have contributed to wide variability in reported sensitivity (ranging from 51% to 100%) and specificity (from 71% to 100%) [9], making cross-study comparisons and synthesis of results challenging. Using data from a Canadian population, our study highlights three key findings: (i) DDSB performance in identifying the presence of SARS-CoV-2 infection was quantified with median sensitivity ranging from 67% to 78% and specificity from 67% to 77% across the three dogs, taking into account the realistic imperfection of RT-PCR; (ii) failure to account for the imperfect nature of both the index test (DDSB) and the reference standard (RT-PCR) led to an underestimation of the DDSB sensitivity, while specificity remained unaffected; and (iii) the inclusion of repeated observations in the analysis, such as samples sniffed more than once by the same dog, without statistical adjustment resulted in an overestimation of the DDSB sensitivity.

Accurate estimation of DDSB performance in identifying COVID-19 is challenged by several potential sources of bias, one of which is the use of various comparison or reference tests (*e.g*., RT-PCR, rapid antigen tests, clinical assessment, CT imaging, and headspace solid-phase microextraction gas chromatography-mass spectrometry [HS-SPME-GC-MS]) without adjusting the analysis for misclassification due to the imperfections of these tests [18,22–39,56–58]. Although RT-PCR is widely used to confirm the diagnosis of COVID-19 in clinical practice [59], it remains an imperfect test due to methodological drawbacks [40–44,60,61]. In addition, the diagnostic performance of RT-PCR for COVID-19 varied considerably across studies, with reported sensitivities ranging from 36.8% to 99.2% and specificities from 79.3% to 99.8% [50]. Performance also varies according to the type of clinical sample tested: sensitivity ranges from 73.6% in serum samples to 100% in respiratory samples, while specificity ranges from 37% in sputum samples to 100% in respiratory, bronchial aspirates, and throat swab samples [62]. Hence, the use of statistical approaches such as Bayesian latent class models is valuable for estimating diagnostic accuracy in the absence of a perfect reference test. These models allow for the estimation of the sensitivity and specificity of the tests being compared, as well as disease prevalence, by treating them as latent parameters and incorporating available prior information [63].

Our study demonstrates that using RT-PCR as the perfect reference test for identifying COVID-19 cases led to a 7.9% to 9.0% underestimation of the posterior median of DDSB sensitivity, without affecting specificity estimates. This finding aligns with the results of Hag-Ali et al. [64], who observed no change in specificity but reported a 6.4% decrease in the estimated sensitivity of DDSB when RT-PCR was assumed to be a perfect reference test. In their study, sensitivity was 89% under the assumption that RT-PCR was an imperfect reference test, compared to 83.3% when the test’s imperfection was not accounted for. Similarly, the study by Guest et al. [65] showed a comparable trend: the sensitivity of human observation of the six included detection dogs decreased by 2.0% to 5.6%, while specificity declined only minimally, by 1.2% to 3.3%, when RT-PCR was treated as the perfect reference test.

Another potential source of bias, the impact of which was assessed in our study, relates to the repeated use of the same samples during validation sessions. Indeed, when a dog is exposed to a sample from an individual it has previously sniffed, either through reuse of the same sample (*e.g*., a training sample or one used in multiple validation sessions) or through a new sample from the same person (as in studies collecting multiple samples per subject, as during our study), its performance may reflect recognition of other volatiles unique to the individual, rather than to the specific disease-related VOCs [49]. In their systematic review of canine olfactory detection of COVID-19, Meller et al. [9] highlighted several studies that repeatedly presented the same samples to the same dogs during validation sessions, without evidence of employing appropriate analytical methods to address this issue. A similar issue may arise when test sessions are recorded and an observer reviews the same video multiple times, leading to repeated coding of the dog’s response to the same sample. Such repeated measurements introduce dependencies between observations, which must be appropriately accounted for in the analysis [66]. In our study, incorporating repeated RT-PCR positive observations into the analysis resulted in higher estimated sensitivity of the DDSB. Another potential source of bias, not assessed in this study due to the small sample size, is the use, during validation sessions, of different samples originating from the same individuals that had already been sniffed by the dogs during training (but different source of sampling).

The observed response of Dog 1, which had no prior olfactory detection experience (“green dog”), showed lower median sensitivity estimates for SARS-CoV-2 infection than those of Dogs 2 and 3, both of which had previous experience in bed bug detection, with differences of approximately 6 to 14 percentage points. However, there was no clear evidence of a statistically meaningful difference, given the substantial overlap of the credible intervals. The influence of prior olfactory detection experience on canine performance has yielded variable results. Some studies suggest that experience with odor detection tasks increases the likelihood of generalization to variations of target odors [67]. In contrast, studies on canine detection of SARS-CoV-2 infection that included both dogs with prior olfactory detection experience (*e.g*., in epilepsy [57] or plant pathogens causing laurel wilt disease [68]) and “green” dogs did not show a performance advantage for experienced dogs. This could also be related to the level of dog expertise influencing the handler. The latter could be more focused on a “green” dog, particularly when working in parallel with more autonomous dogs, or at the inverse be less patient with a “green” dog.

Although our study highlights the significant impact of a few specific biases on the estimation of DDSB performance, diagnostic studies involving dogs are subject to multiple sources of bias. These biases may arise at various stages of the study, such as sampling, sample preparation, storage and handling, training and validation phases, and data analysis, and may involve a range of stakeholders, including participants, the research team, clinicians, laboratory staff, dogs, handlers, observers, and data analysts. While standardizing practices across studies and operational contexts presents significant challenges, an important first step is to adhere to existing guidelines [49], thoroughly document study procedures to enable critical evaluation, and apply appropriate statistical methods for data analysis.

This study has several limitations. First, we encountered difficulties in recruiting a qualified dog handler, which contributed to delays between the collection of samples and their use in dog training. Additionally, since the samples were used 4 to 6 months after collection, it is difficult to assess the extent to which the quality of VOCs may have deteriorated, potentially affecting the dogs’ performance. The validation phase did not proceed as planned, as sessions were prematurely terminated due to unforeseen circumstances, thereby explaining the discrepancy between the number of participants initially recruited and those ultimately included in the validation phase. It can be challenging for some handlers to be kept blinded to the type of samples sniffed by the dogs as they may feel judged by the performance of their dogs. However, the sample included in the short testing phase which did take place were selected randomly from the whole population sampled. Furthermore, the unexpected interruption of the validation meant that we could not estimate between-dog and within-dog variability. To adjust for repetition *via* modeling, we would have needed a balanced design with a similar number of repetitions for each subject. As previously noted, the analyzed data included samples assigned to the validation from some individuals whose other samples had previously been used during the training phase, as well as some positive samples that were sniffed more than once by the dogs. The latter were addressed in the analysis by excluding duplicate observations, whereas the former could not be accounted for, due to the limited sample size available. Finally, the conditions in which the dogs were trained and tested were not ideal since it was not done in an existing detection dog center with already available and familiar material and equipment. In addition, the equipment used to present the samples to the dogs changed between steps 4 and 5 of the training. These elements could have influenced the performance of the dogs due to the stress related to a new environment and equipment. Despite all of this, the median values of the estimated sensitivity and specificity were considered as encouraging.

In conclusion, our study showed that DDSB demonstrated promising performance as a non-invasive screening tool for SARS-CoV-2 infection. However, sensitivity estimates were influenced by selected sources of bias, including repeated sample sniffing by the dogs and the failure to account for the imperfect accuracy of RT-PCR as the reference standard. The current gold standard in screening for COVID-19 presents challenges of invasive testing, cost, timing, personnel required, and diagnostic accuracy. There is a pressing need for a rapid, noninvasive and sensitive means for screening large populations. With multiple studies reporting the effectiveness of detection dogs in detecting various human diseases, there is growing interest in the field deployment of trained dogs as reliable frontline clinical screening tools. Deploying the use of trained detection dogs could be an effective and rapid diagnostic tool to screen people for COVID-19 or other communicable diseases in various settings of public spaces, mass gatherings, outbreak settings, and for at-risk countries, particularly in underserved or remote areas with limited access to healthcare services. However, questions remain regarding the heterogeneity in accuracy reported across studies. Moreover, it remains to be studied whether the observed accuracy translates into rapid, cost-effective testing equivalent to available clinical tests. To achieve this goal, the formal standardization of training and validation protocols, along with an assessment of feasibility in practical settings, is essential and further studies are needed [45,59].

## Data Availability

All relevant data are within the manuscript and its Supporting Information files.

## Acknowledgments

The authors would like to thank Mr. Yassar Muttaqui of the Ontario Ministry of Colleges and Universities, Research and Innovation Division, Mrs. Stacie Korjenevitch, executive assistant to Member of Provincial (Ontario) Parliament (MPP) Mrs. Kinga Surma, MPP for Etobicoke Centre, and Mrs. Kinga Surma herself for their incredible support, Prof. Dominique Grandjean of the Nosaïs Institute / École Nationale Vétérinaire d’Alfort, Paris – France, and Prof. Anna Hielm-Björkman of the DogRisk Research Group / University of Helsinki, Finland, the Unity Health Ontario for their so efficient partnership and advices, the Conseil Scolaire Viamonde for their collaboration in transforming a school in a canine center, Mr. Emmanuel Klein and Mr. Damien Truffaut of Sixt Sense, Inc. for the video-analysis system, Dr. Nickolay Chirgadze of X-CHIP Technologies, Inc. for the sniffing station, Mr. David Jeanneault from Frontline K9 Consulting LP, and Mr Mario Lavigne for his enthusiasm and interest in dog detection.

## Supporting information

**S1 Appendix. STARD-BLCM Checklist.**

**S2 Appendix. Models.**

**S1 Table. Diagnostic accuracy of human observation of three trained detection dogs in identifying COVID-19 compared to RT-PCR as the reference test.**

**S2 Table. Diagnostic accuracy of RT-PCR when assumed to be an imperfect reference test.**

**S3 Table. Cross-tabulation of the test results.**

