## Supplementary material for "Assessment of accuracy of detection dog signaling behavior for the diagnosis of SARS-CoV-2 infection: A Canadian study": S1 Appendix

**STARD-BLCM Checklist**

This checklist was adapted from the model proposed by Kostoulas et al.: STARD-BLCM: Standards for the Reporting of Diagnostic accuracy studies that use Bayesian Latent Class Models. Prev Vet Med. 2017;138:37-47.

| **Section & topic** | **Item** | **STARD-BLCM** | **Reported on page #** |
| --- | --- | --- | --- |
| Title/Abstract/Keywords | 1 | Identification as a study of diagnostic accuracy, using at least one measure of accuracy (such as sensitivity, specificity, predictive values, or AUC) and Bayesian latent class models | 1, 3–4 |
| Abstract | 2 | Structured summary of study design, methods, results, and conclusions (for specific guidance, see STARD for Abstracts) | 3–4 |
| Introduction | 3 | Scientific and clinical background, including the intended use and clinical role of the tests under evaluation | 4–6 |
|  | 4 | Study objectives and hypotheses, such as estimation of diagnostic accuracy of the tests for a defined purpose through BLCM | 6 |
| **Methods** | | |  |
| *Study Design* | 5 | Whether data collection was planned before the tests were performed (prospective study) or after (retrospective study) | 6–12 |
| *Participants* | 6 | Eligibility criteria and description of the source population | 7 |
|  | 7 | On what basis potentially eligible participants were identified (such as symptoms, results from previous tests, inclusion in registry) | 7 |
|  | 8 | Where and when potentially eligible participants were identified (setting, location, and dates) | 7 |
|  | 9 | Whether participants formed a consecutive, random or convenience series | 7 |
| *Test methods* | 10 | Description of the tests under evaluation, in sufficient detail to allow replication, and/or cite references | 8–13 |
|  | 11 | Rationale for choosing the tests under evaluation in relation to their purpose | 4–6 |
|  | 12 | Rationale for test positivity cut-offs or result categories of the tests under evaluation, distinguishing pre-specified from exploratory | 8–10 |
|  | 13 | Whether clinical information was available to the performers or readers of the tests under evaluation | N/A |
| *Analysis* | 14a | BLCM model for estimating measures of diagnostic accuracy | 13 |
|  | 14b | Definition and rationale of prior information and sensitivity analysis | 13 |
|  | 15 | How indeterminate results of the tests under evaluation were handled | Fig 2; S1 and S3 Tables |
|  | 16 | How missing data of the tests under evaluation were handled | N/A |
|  | 17 | Any analyses of variability in diagnostic accuracy, distinguishing pre-specified from exploratory | N/A |
|  | 18 | Intended sample size and how it was determined | N/A |
| **Results** | | |  |
| *Participants* | 19 | Flow of participants, using a diagram | Fig 2 |
|  | 20 | Baseline demographic and clinical characteristics of participants | 14–15 |
|  | 21a | Distribution of severity of disease in those with the target condition | N/A |
|  | 21b | Distribution of alternative diagnoses in those without the target condition | N/A |
|  | 22 | Time interval and any clinical interventions between the tests under evaluation | 7–9 |
| *Test results* | 23 | Cross tabulation of the tests’ results (or for continuous tests results their distribution by infection stage) | S3 Table |
|  | 24 | Estimates of diagnostic accuracy under alternative prior specification and their precision (such as 95% credible/probability intervals) | 16–18; S1 and S2 Tables |
|  | 25 | Report any adverse events from performing the of the tests under evaluation | N/A |
| **Discussion** | | | |
|  | 26 | Study limitations, including sources of potential bias, statistical uncertainty, and generalisability | 18–22 |
|  | 27 | Implications for practice, including the intended use and clinical role of the tests under evaluation in relevant settings (clinical, research, surveillance etc.) | 22–23 |
| **Other information** | | |  |
|  | 28 | Registration number and name of registry | N/A |
|  | 29 | Where the full study protocol can be accessed | N/A |
|  | 30 | Sources of funding and other support; role of funders | N/A  (statement added in final publication by publisher) |
