## Supplementary material for "Assessment of accuracy of detection dog signaling behavior for the diagnosis of SARS-CoV-2 infection: A Canadian study": S2 Appendix

**Models**

1. **JAGS Model for Estimating the Sensitivity and Specificity of DDSB Assuming Reverse Transcription Polymerase Chain Reaction (RT-PCR) as a Perfect Reference Test**

model {

for (i in 1:N) {

#============

\# LIKELIHOOD

#============

D1[i]~dbern(prev)

D[i]<-D1[i]+1

\# PCR

y[i,1]~dbern(p1[i,D[i]])

\# Conditional probability of a positive observation

p1[i,2]<-s1[i]

p1[i,1]<-1-c1[i]

s1[i]<-1

c1[i]<-1

\# DDSB "TEST"

y[i,2]~dbern(p2[i,D[i]])

\# Conditional probability of a positive observation

p2[i,2]<-s2[i]

p2[i,1]<-1-c2[i]

s2[i]<-phi(a[2,1])

c2[i]<-phi(a[2,2])

}

a[2,1]<-probit(se[2])

a[2,2]<-probit(sp[2])

#==================================================

\# Prior distributions

#==================================================

prev~dbeta(1,1)

se[1]<-1

sp[1]<-1

se[2]~dbeta(1,1)

sp[2]~dbeta(1,1)

\# ACCURACY FOR PCR

S.OVERALL[1] <- 1

C.OVERALL[1] <- 1

\# ACCURACY FOR DDSB

S.OVERALL[2] <- mean(s2[])

C.OVERALL[2] <- mean(c2[])

S <- phi(a[2,1])

C <- phi(a[2,2])

}

2. **JAGS Model for Estimating the Sensitivity and Specificity of DDSB Assuming Reverse Transcription Polymerase Chain Reaction (RT-PCR) as an imperfect Reference Test**

model {

for (i in 1:N) {

#============

\# LIKELIHOOD

#============

D1[i]~dbern(prev)

D[i]<-D1[i]+1

\# PCR

y[i,1]~dbern(p1[i,D[i]])

\# Conditional probability of a positive observation

p1[i,2]<-s1[i]

p1[i,1]<-1-c1[i]

s1[i]<-phi(a[1,1])

c1[i]<-phi(a[1,2])

\# DDSB "TEST"

y[i,2]~dbern(p2[i,D[i]])

\# Conditional probability of a positive observation

p2[i,2]<-s2[i]

p2[i,1]<-1-c2[i]

s2[i]<-phi(a[2,1])

c2[i]<-phi(a[2,2])

}

a[1,1] <- probit(se[1])

a[1,2] <- probit(sp[1])

a[2,1]<-probit(se[2])

a[2,2]<-probit(sp[2])

#==================================================

\# Prior distributions

#==================================================

prev~dbeta(1,1)

\# Informative prior for PCR test derived from the systematic review by Vilca-Alosilla, J.J.; Candia-Puma, M.A.; Coronel-Monje, K.; Goyzueta-Mamani, L.D.; Galdino, A.S.; Machado-de-Ávila, R.A.; Giunchetti, R.C.; Ferraz Coelho, E.A.; Chávez-Fumagalli, M.A. A Systematic Review and Meta-Analysis Comparing the Diagnostic Accuracy Tests of COVID-19. Diagnostics 2023, 13, 1549. https://doi.org/10.3390/diagnostics13091549

se[1]~dbeta(53.1006251, 4.0260205)

sp[1]~dbeta(29.2364884, 1.2467747)

se[2]~dbeta(1,1)

sp[2]~dbeta(1,1)

\# ACCURACY FOR PCR

S.OVERALL[1] <- mean(s1[])

C.OVERALL[1] <- mean(c1[])

\# ACCURACY FOR DDSB

S.OVERALL[2] <- mean(s2[])

C.OVERALL[2] <- mean(c2[])

S[1] <- phi(a[1,1])

S[2] <- phi(a[2,1])

C[1] <- phi(a[1,2])

C[2] <- phi(a[2,2])

}
