## Supplementary material for "Assessment of accuracy of detection dog signaling behavior for the diagnosis of SARS-CoV-2 infection: A Canadian study": S1 Table

**S1A Table. Diagnostic accuracy of human observation of three trained detection dogs in identifying COVID-19 compared to RT-PCR as the reference test. Unique participation of sample per dog.**

|  | **Dog 1** | **Dog 2** | **Dog 3** |
| --- | --- | --- | --- |
| Number of samples examined | 52 | 70/71^a^ | 60 |
|  | **RT-PCR assumed perfect** | | |
| Sensitivity (%), median (95% CrI) |  |  |  |
| Observer 1 | 61 (30, 86) | 71 (40, 92) | 70 (43, 90) |
| Observer 2 | 61 (30, 86) | 74 (44, 93) | 70 (43, 90) |
| Specificity (%), median (95% CrI) |  |  |  |
| Observer 1 | 74 (61, 85) | 77 (65, 86) | 67 (53, 79) |
| Observer 2 | 74 (61, 85) | 86 (77, 93) | 71 (58, 82) |
|  | **RT-PCR assumed imperfect** | | |
| Sensitivity (%), median (95% CrI) |  |  |  |
| Observer 1 | 67 (29, 97) | 78 (41, 99) | 76 (44, 98) |
| Observer 2 | 67 (29, 97) | 81 (48, 99) | 76 (44, 98) |
| Specificity (%), median (95% CrI) |  |  |  |
| Observer 1 | 74 (60, 86) | 77 (65, 87) | 67 (53, 79) |
| Observer 2 | 74 (60, 86) | 87 (77, 94) | 71 (58, 83) |

RT-PCR, reverse transcription-polymerase chain reaction; CrI, credible interval.

^a^ The total number of samples was 70 for observer 1 and 71 for observer 2, as one sample was excluded from the analysis involving observer 1 due to an inconclusive result reported for Dog 2.

**S1B Table. Diagnostic accuracy of human observation of three trained detection dogs in identifying COVID-19 compared to RT-PCR as the reference test. Including same samples repeated in testing per dog.**

|  | **Dog 1** | **Dog 2** | **Dog 3** |
| --- | --- | --- | --- |
| Number of samples examined | 55 | 77/78^a^ | 64 |
|  | **RT-PCR assumed perfect** | | |
| Sensitivity (%), median (95% CrI) |  |  |  |
| Observer 1 | 70 (42,90) | 84 (62, 96) | 78 (54, 93) |
| Observer 2 | 70 (43, 90) | 85 (64, 96) | 78 (54, 93) |
| Specificity (%), median (95% CrI) |  |  |  |
| Observer 1 | 74 (61, 85) | 77 (65, 86) | 67 (53, 79) |
| Observer 2 | 74 (61, 85) | 86 (77, 93) | 71 (58, 82) |
|  | **RT-PCR assumed imperfect** | | |
| Sensitivity (%), median (95% CrI) |  |  |  |
| Observer 1 | 76 (45, 98) | 88 (64, 99) | 82 (57, 99) |
| Observer 2 | 76 (44, 98) | 89 (67, 99) | 83 (56, 98) |
| Specificity (%), median (95% CrI) |  |  |  |
| Observer 1 | 75 (61, 87) | 78 (66, 88) | 68 (54, 80) |
| Observer 2 | 75 (61, 87) | 88 (78, 95) | 72 (58, 84) |

RT-PCR, reverse transcription-polymerase chain reaction; CrI, credible interval.

^a^ The total number of samples was 77 for observer 1 and 78 for observer 2, as one sample was excluded from the analysis involving observer 1 due to an inconclusive result reported for Dog 2.
