## Supplementary material for "Assessment of accuracy of detection dog signaling behavior for the diagnosis of SARS-CoV-2 infection: A Canadian study": S2 Table

**S2 Table. Diagnostic accuracy of RT-PCR when assumed to be an imperfect reference test.**

|  | **Dog 1** | **Dog 2** | **Dog 3** |
| --- | --- | --- | --- |
| Sensitivity (%), median (95% CrI) | 93 (85, 99) | 93 (85, 98) | 93 (85, 98) |
| Specificity (%), median (95% CrI) | 97 (89, 100) | 98 (92, 100) | 97 (89, 100) |

CrI, credible interval.
