## Supplementary material for "Assessment of accuracy of detection dog signaling behavior for the diagnosis of SARS-CoV-2 infection: A Canadian study": S3 Table

**S3A Table. Cross-tabulation of the test results for each dog. Unique participation of sample per dog.**

| **Observer #** | **Dog #** | **RT-PCR + and DDSB +** | **RT-PCR - and DDSB +** | **RT-PCR + and DDSB -** | **RT-PCR - and DDSB -** | **Total** |
| --- | --- | --- | --- | --- | --- | --- |
| Observer 1 | 1 | 5 | 11 | 3 | 33 | 52 |
|  | 2 | 6 | 14 | 2 | 48 | 70^a^ |
|  | 3 | 8 | 16 | 3 | 33 | 60 |
| Observer 2 | 1 | 5 | 11 | 3 | 33 | 52 |
|  | 2 | 7 | 8 | 2 | 54 | 71^a^ |
|  | 3 | 8 | 14 | 3 | 35 | 60 |

RT-PCR, reverse transcription-polymerase chain reaction; DDSB, Detection Dog Signaling Behavior.

^a^ The total number of samples was 70 for observer 1 and 71 for observer 2, as one sample was excluded from the analysis involving observer 1 due to an inconclusive result reported for Dog 2.

**S3B Table. Cross-tabulation of the test results for each dog. Including same samples repeated in testing per dog.**

| **Observer #** | **Dog #** | **RT-PCR + and DDSB +** | **RT-PCR - and DDSB +** | **RT-PCR + and DDSB -** | **RT-PCR - and DDSB -** | **Total** |
| --- | --- | --- | --- | --- | --- | --- |
| Observer 1 | 1 | 8 | 11 | 3 | 33 | 55 |
|  | 2 | 13 | 14 | 2 | 48 | 77^a^ |
|  | 3 | 12 | 16 | 3 | 33 | 64 |
| Observer 2 | 1 | 8 | 11 | 3 | 33 | 55 |
|  | 2 | 14 | 8 | 2 | 54 | 78^a^ |
|  | 3 | 12 | 14 | 3 | 35 | 64 |

RT-PCR, reverse transcription-polymerase chain reaction; DDSB, Detection Dog Signaling Behavior.

^a^ The total number of samples was 77 for observer 1 and 78 for observer 2, as one sample was excluded from the analysis involving observer 1 due to an inconclusive result reported for Dog 2.
